# Clinical Laboratory Parameters Associated with Severe or Critical Novel Coronavirus Disease 2019 (COVID-19): A Systematic Review and Meta-analysis

**DOI:** 10.1101/2020.04.24.20078782

**Authors:** Jude Moutchia, Pratik Pokharel, Aldiona Kerri, Kaodi McGaw, Shreeshti Uchai, Miriam Nji, Michael Goodman

## Abstract

**Background:** To date, several clinical laboratory parameters associated with COVID-19 severity have been reported. However, these parameters have not been observed consistently across studies. The aim of this review was to assess clinical laboratory parameters which may serve as markers or predictors of severe or critical COVID-19 disease

**Methods:** We conducted a systematic search of MEDLINE, Embase, Web of Science, CINAHL and Google Scholar databases from 2019 through April 18, 2020, and reviewed bibliographies of eligible studies, relevant systematic reviews, and the medRxiv pre-print server. We included hospital-based observational studies reporting clinical laboratory parameters of confirmed cases of COVID-19 and excluded studies having large proportions (>10%) of children and pregnant women. Two authors independently carried out screening of articles, data extraction and quality assessment. Meta-analyses were done using random effects model. Meta-median difference (MMD) and 95% confidence interval (CI) was calculated for each laboratory parameter.

**Results:** Forty-five studies in 6 countries were included. Compared to non-severe COVID-19 cases, severe or critical COVID-19 disease was characterised by higher neutrophil count (MMD: 1.23 [95% CI: 0.58 to 1.88] ×10^9^ cells/L), and lower lymphocyte and CD4 counts with MMD (95% CI) of -0.39 (-0.47, -0.31) ×10^9^ cells/L and -204.9 (-302.6, -107.1) cells/μl, respectively. Other notable results were observed for C-reactive protein (MMD: 36.97 [95% CI: 27.58, 46.35] mg/L), interleukin-6 (MMD: 17.37 [95% CI: 4.74, 30.00] pg/ml,), Troponin I (MMD: 0.01 [0.00, 0.02] ng/ml), and D-dimer (MMD: 0.65 [0.45, 0.85] mg/ml).

**Conclusions and Relevance:** Relative to non-severe COVID-19, severe or critical COVID-19 is characterised by increased markers of innate immune response, decreased markers of adaptive immune response, and increased markers of tissue damage and major organ failure. These markers could be used to recognise severe or critical disease and to monitor clinical course of COVID-19.

## Introduction

Coronavirus disease 2019 (COVID-19) is an emerging zoonosis caused by Severe Acute Respiratory Syndrome Coronavirus 2 (SARS-CoV-2) [1, 2]. Phylogenetically, SARS-CoV-2 sufficiently differs from other zoonotic coronaviruses, such as Severe Acute Respiratory Syndrome Coronavirus (SARS-CoV) and Middle East Respiratory Syndrome Coronavirus (MERS-CoV) introduced to humans in the past two decades [1, 3]. Disease resulting from infection with SARS-CoV-2 was first reported in Wuhan, China in December 2019, and the virus rapidly spread to other regions of the world thereafter [4, 5]. Given the scale of the outbreak, COVID-19 was declared a pandemic on March 12 2020 by the World Health Organization [6]. As of April 19, 2020, there have been 2,394,291 confirmed cases in 185 countries/regions and 164,938 COVID-related deaths [7].

Clinical features of infection with SARS-CoV-2 vary widely and have been classified as mild, severe or critical, with some persons remaining asymptomatic [8, 9]. Majority of SARS-CoV-2 infected persons display mild symptoms similar to a viral upper respiratory tract infection such as dry cough, fever, sore throat, nasal congestion, and muscle pain [8-10]. Severe COVID-19 is characterised by features of severe pneumonia such as dyspnoea, respiratory frequency ≥30 breaths per minute and blood oxygen saturation ≤93%, while critical COVID-19 is characterised by respiratory failure, septic shock, and/or multiple organ failure [8, 9]. Severe or critical COVID-19 is highly associated with mortality [11]. In a single-centre observational study of critical COVID-19 patients, up to 61% of critical COVID-19 patients and 94% of critical COVID-19 patients requiring mechanical ventilation died within 28 days of admission into the intensive care unit [12].

Currently, there is no approved cure for infection with SARS-CoV-2 and an effective vaccine is not yet available. Approximately 18% of diagnosed COVID-19 cases have severe or critical disease, and about 5% of diagnosed COVID-19 require intensive care management with or without mechanical ventilation [8, 13]. Consequently, there is substantial pressure on healthcare systems worldwide, particularly on intensive care units. As healthcare systems become further stretched by the increasing numbers of cases, identifying clinical laboratory parameters associated with severe and critical cases is crucial in helping clinicians triage patients appropriately and optimize use of the limited healthcare resources. Furthermore, as more clinical trials are being launched to test possible treatments for COVID-19, laboratory parameters associated with COVID-19 severity can aid in monitoring the clinical evolution of cases on trial drugs and serve as composite or secondary outcomes for these trials.

To date, changes in several clinical laboratory parameters have been linked to COVID-19 severity [4, 13-16]. However, it is not clear if these changes are observed consistently across studies. With these considerations in mind, the objective of this systematic review and meta-analysis was to investigate which clinical laboratory parameters may be associated with severe or critical COVID-19 disease.

## Methods

### Protocol and registration

We registered our study protocol with the International Prospective Register of Systematic Reviews (PROSPERO); registration number CRD42020176651 [17]. This review and meta-analysis was conducted and has been reported according to The Preferred Reporting Items for Systematic Reviews and Meta-Analyses (PRISMA) statement and Meta-analysis of Observational Studies in Epidemiology (MOOSE) guidelines [18, 19].

### Eligibility criteria

This review and meta-analysis included observational studies reporting clinical laboratory parameters among patients with confirmed COVID-19. Cases were diagnosed using guidelines by either the World Health Organization or the China National Commission for Health [20, 21].

The exposure of interest of this review was severe or critical COVID-19 and the comparator was non-severe COVID-19. According to the criteria defined by China National Health Commission, severe COVID-19 is characterised by dyspnoea, ≥30 breaths/minute, blood oxygen saturation ≤93%, arterial partial pressure of oxygen to fraction of inspired oxygen (PaO^2^/FiO^2^) ratio <300, and/or lung infiltrates >50% within 24–48 hours; and critical COVID-19 is characterised by respiratory failure, septic shock, and/or multiple organ failure [22]. Non-severe COVID-19 is defined by no or mild pneumonia [22]. We also considered COVID-19 cases requiring oxygen therapy, and COVID-19 cases admitted to intensive care units as severe or critical cases.

The outcomes of interest were clinical laboratory parameters. These included hematologic indices (White blood cells, Neutrophils, Lymphocytes, Monocytes, Platelets, Haemoglobin, CD3, CD4, CD8), biochemical indices (Total bilirubin, Alanine aminotransferase, Aspartate aminotransferase, Total protein, Albumin, Globulin, Prealbumin, Urea, Creatinine, Glucose, Creatine kinase muscle-brain, Troponin I, Cholinesterase, Cystatin C, Lactate dehydrogenase, α-hydroxybutyric dehydrogenase), infection/inflammation-related indices (C-reactive protein [high sensitivity and standard], Interleukin-6, Erythrocyte sedimentation rate, Procalcitonin, Serum ferritin), coagulation indices (Prothrombin time, Activated partial thromboplastin, D-dimer) and electrolytes (Sodium, Potassium, Calcium, Chloride).

We included only hospital-based studies and excluded reviews, opinion articles, and studies that did not report clinical laboratory parameters stratified by COVID-19 disease severity. Also, as children and pregnant women have different cut-off values for most clinical laboratory parameters compared to general adults, we excluded studies that examined populations with large proportions of children under 11 years of age and pregnant women to reduce clinical heterogeneity. We considered studies that included children, pregnant women along with the general adult population as eligible only if the proportion of children or pregnant women constituted less than 10%.

### Search strategy

We conducted a systematic search of Ovid MEDLINE, Ovid Embase, Clarivate Analytics Web of Science Core Collection, EBSCO CINAHL and Google Scholar databases from 2019 through April 18, 2020. The search strategy used both controlled vocabulary and free text words relevant to COVID-19 and clinical laboratory parameters (see search strategy in Supplement S1). We also reviewed bibliographies of eligible studies, relevant systematic reviews to identify additional papers that were missed by the electronic search. Further, we performed a manual search of the medRxiv pre-print server to identify latest relevant studies that might still be undergoing peer-review. The search was limited to the years 2019-2020 and there was no limitation regarding language of publication.

### Study selection

Following deduplication of records retrieved during the systematic search, we exported retained articles into Covidence review manager to facilitate the screening of titles and abstracts, which was followed by a full text review to determine eligibility [23].

Two authors (JM and PP) independently carried out title and abstract screening and full text evaluation of all articles using the eligibility criteria listed in the previous section. The discrepancies in study selection were resolved through adjudication by a third author (KM). To avoid including data on the same patient populations more than once in the meta-analysis, we matched studies based on the location of the study (hospital, town) and the period over which data was collected. For two or more studies conducted at the same location over the same or overlapping periods, we included only the largest study, unless one of the smaller studies presented relevant information not included in the larger study.

### Data extraction and Data items

Two authors (JM and PP) independently extracted, verified and summarized data from each study included in the meta-analysis. The information extracted from the selected studies included: study author(s), study sponsors, date of publication, study period, study location, study design, sample size, sample characteristics (age, gender, comorbidities), exposure characteristics (study definition of severity of COVID-19, timing of classification of disease severity [on admission or otherwise], number of cases with non-severe COVID-19, number of cases with severe or critical COVID-19), timing of blood sample collection (on admission or otherwise), clinical laboratory parameters stratified by COVID-19 severity, mean (standard deviation [SD]) and/or median (interquartile range [IQR] or minimum-maximum [total] range) of clinical laboratory parameters when reported on continuous scales, and numbers (percentages) of cases above and below cut-off values when reported on categorical scales. Discrepancies in collected data were resolved by re-checking the primary studies until consensus was reached. For the studies which had unclear severity classification, the authors were contacted to seek additional clarification. Studies in the Chinese language were translated into English language by a Chinese native speaker. The extracted data were exported into R programming software.

### Quality assessment

Two authors (AK and MN) independently carried out quality assessment of each article using National Institutes of Health (NIH) study quality assessment tools for observational cohort and cross-sectional studies, and for case series studies [24]. These tools were used to evaluate the risk of bias and to assess the overall validity of reported results. Each study was assessed using all elements of the relevant tool, and an overall judgement was made by considering the responses to the various elements. An overall rating of poor quality translates to a high risk of bias, and an overall rating of good quality translates to a low risk of bias [24]. The final decision for each study was made through professional judgement and by consensus among the authors. We evaluated the impact of studies with a high risk of bias by doing sensitivity analysis using the Leave-One-Out method [25].

### Summary measures and data synthesis

Where clinical laboratory parameters were measured on a continuous scale, we pooled median differences from each study using the quantile estimation method [26]. The result of this analysis was expressed as a meta-median difference (MMD) accompanied by a corresponding 95 % confidence interval (CI). We preferred median differences over mean differences because clinical laboratory indices are usually skewed, and mean values could be influenced by outlier values, particularly in small samples. We performed a sensitivity analysis by pooling mean differences from each study using inverse variance weighting. Where the studies reported only median (IQR or total range) values, we computed mean (SD) using methods previously described [27, 28].

Where clinical laboratory parameters were measured on a categorical scale, we computed prevalence ratios for each study using counts of events in the exposure and comparator group and calculated meta-prevalence ratios (MPR) and the 95% CIs using the Mantel-Haenszel method.

Meta-analysis was conducted using random effect models. We assessed clinical heterogeneity (age distribution, comorbidities criteria of severity) and study methodological heterogeneity (timing of blood sample collection) and considered the potential impact of these factors on the meta-analysis results. We assessed statistical heterogeneity using Cochran’s Q test and calculated the I^2^ statistic, which was interpreted using cut-offs of 25%, 50%, and 75% for low, moderate, and substantial heterogeneity, respectively. We performed influence analysis using the Leave-One-Out-method to identify studies that have a high influence on our results [25]. Additional sensitivity analyses were performed by excluding ‘outlier’ studies. A study-specific estimate was considered an outlier if its confidence interval did not overlap with the confidence interval of the meta-estimate [25].

To detect possible publication bias, funnel plots were constructed for the 4 laboratory parameters with the highest number of individual studies. Egger’s test was carried out to assess statistical symmetry of the plots.

Statistical analyses were done using R programming software and in the ‘meta’, ‘metafor’, ‘dmetar’ and ‘metadian’ packages [29].

## Results

### Study selection

We identified 3,779 studies through database searching and from other sources (Figure 1). After removing duplicates, 1722 unique records were screened, and of those, 1398 were removed after title and abstract review. Additional 257 records were excluded due to lack of COVID-19 severity classification, lack of laboratory parameter records or ineligible study design. Of the 67 remaining studies, another 22 were excluded because they used data from the same locations or covered overlapping periods (see Supplement S2). A total of 45 studies were retained for meta-analyses.

**Figure 1:**
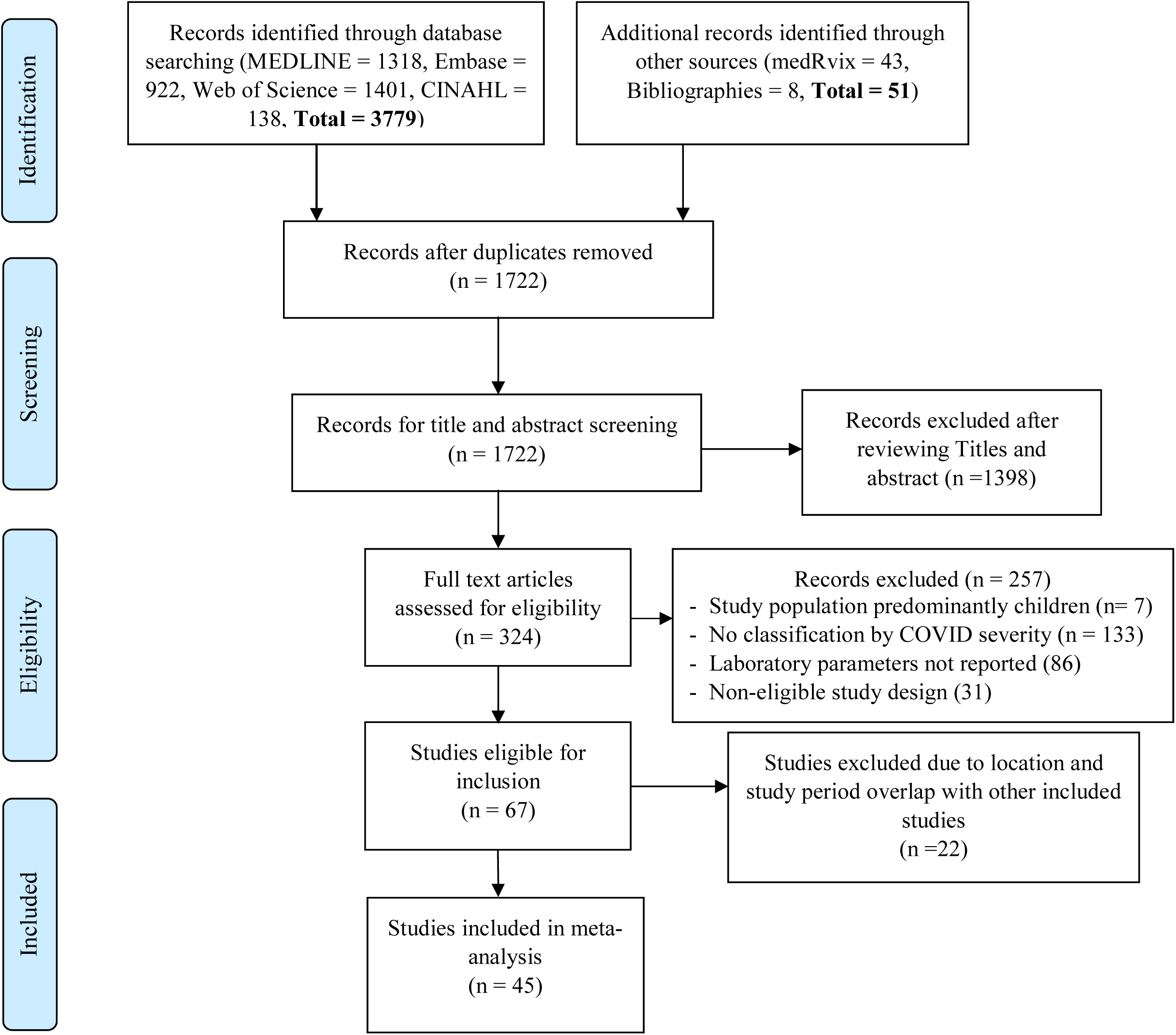
PRISMA flowchart of study selection

### Study characteristics

The characteristics of included studies are detailed in Table 1 and Supplement S3. All studies included in the meta-analyses were observational and hospital based. The majority of 45 studies (87%) were from China; and of those, 14 were from Wuhan and 25 from other locations in China. Two studies were from the USA, and the remaining 4 studies were from France, Germany, Japan and Singapore. All studies were published in 2020 and the data collection covered the period from December 25^th^, 2019 to April 2^nd^, 2020. The median population size of the included studies was 97 (IQR: 49 – 221). Data were collected retrospectively in all but one study [30]. COVID-19 severity was classified using China National Health Commission guidelines (20 studies), WHO guidelines (4 studies), American Thoracic Society guidelines (2 studies), Berlin criteria (1 study), Complementary and Natural Healthcare council (1 study), or unspecified guidelines (17 studies). Studies classified severity on admission (22 studies, 49%), on the ward (11 studies, 24%), or during unspecified periods (12 studies, 27%). Clinical laboratory tests were done on admission (33 studies, 73%), post-admission (5 studies, 11%) and at unspecified periods (7 studies, 16%). The highest number of laboratory parameters reported in a single study was 30 [31] and the lowest number of laboratory parameters reported in a single study was 2 [32].

**Table 1:**
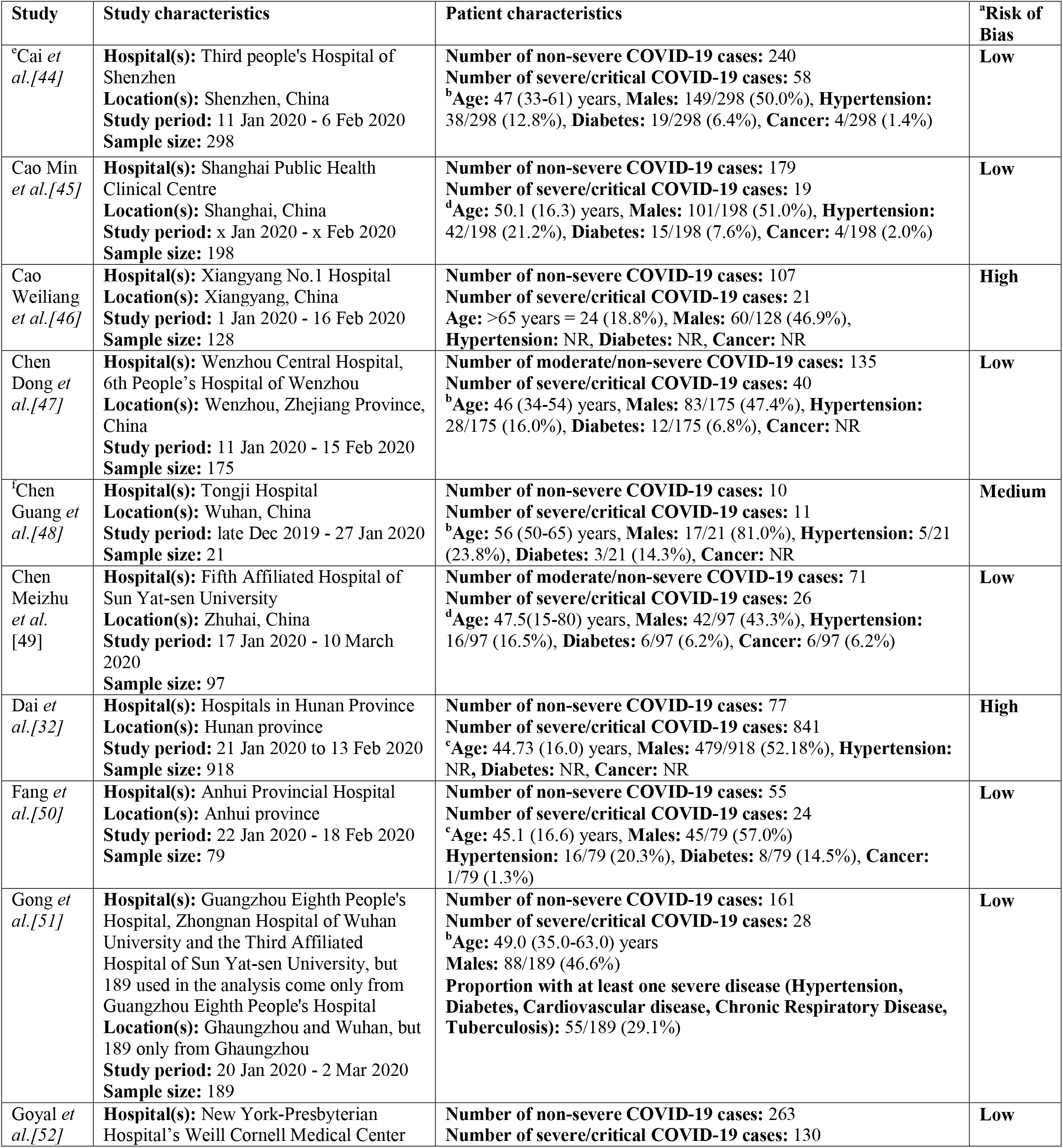
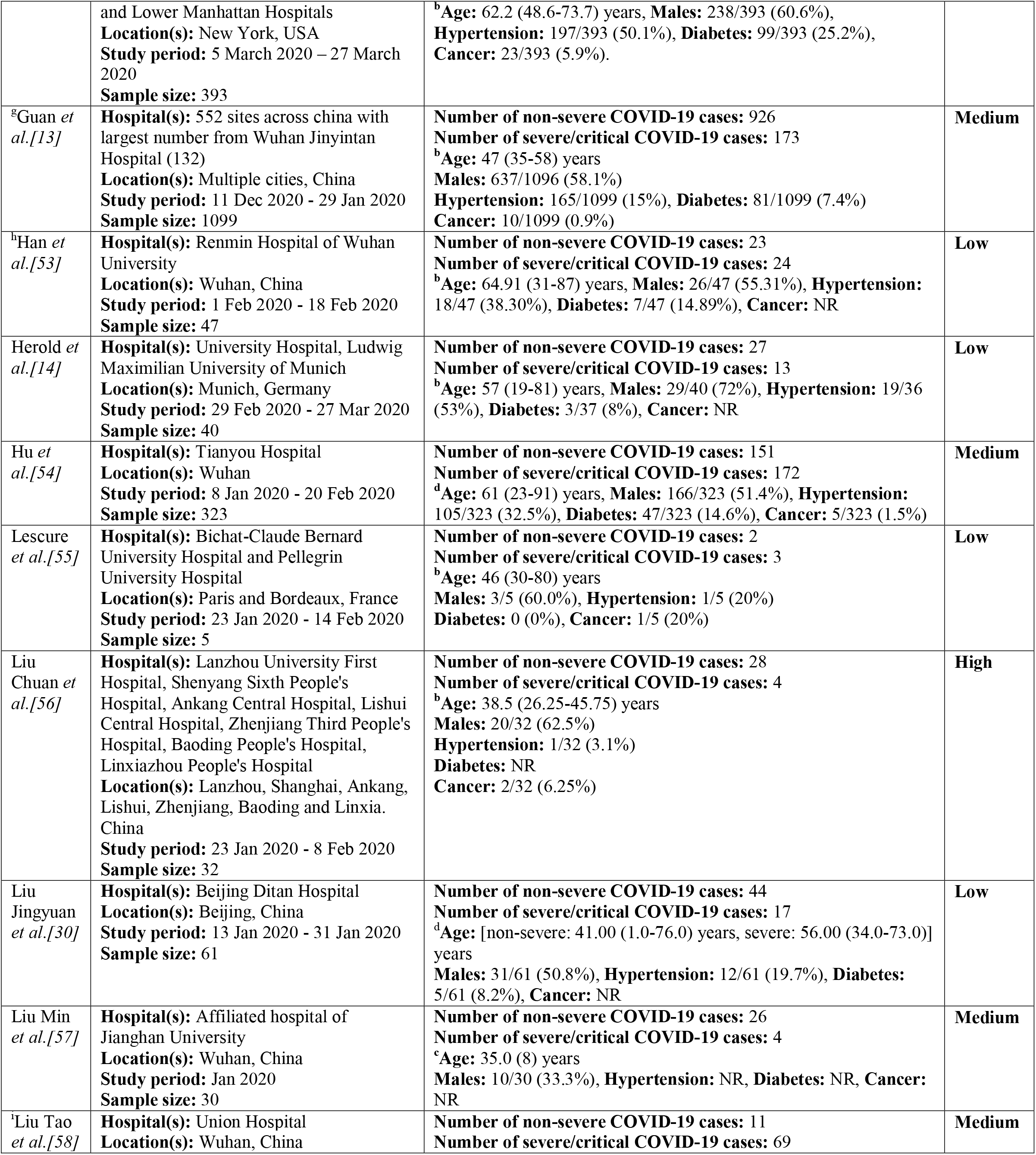
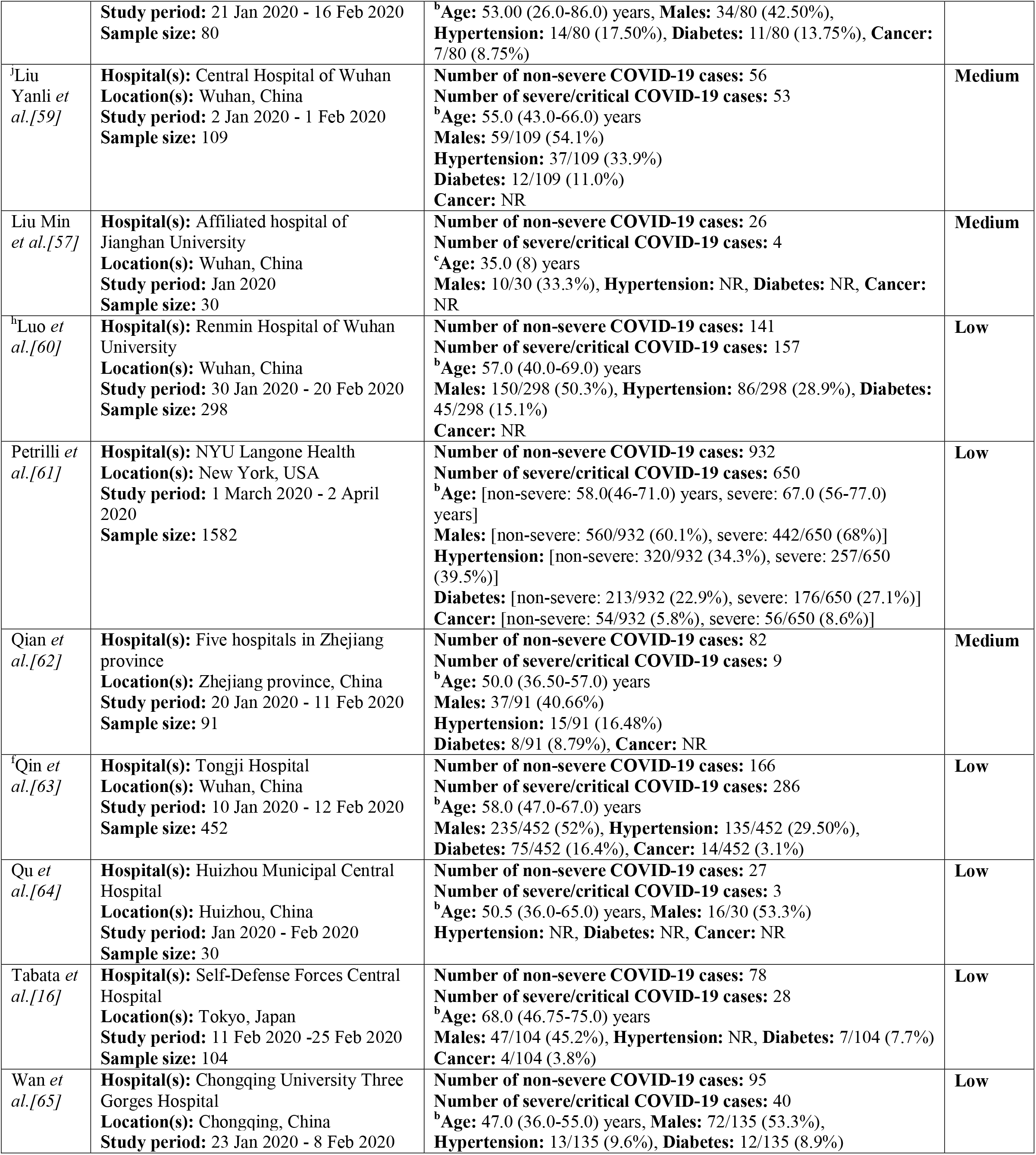
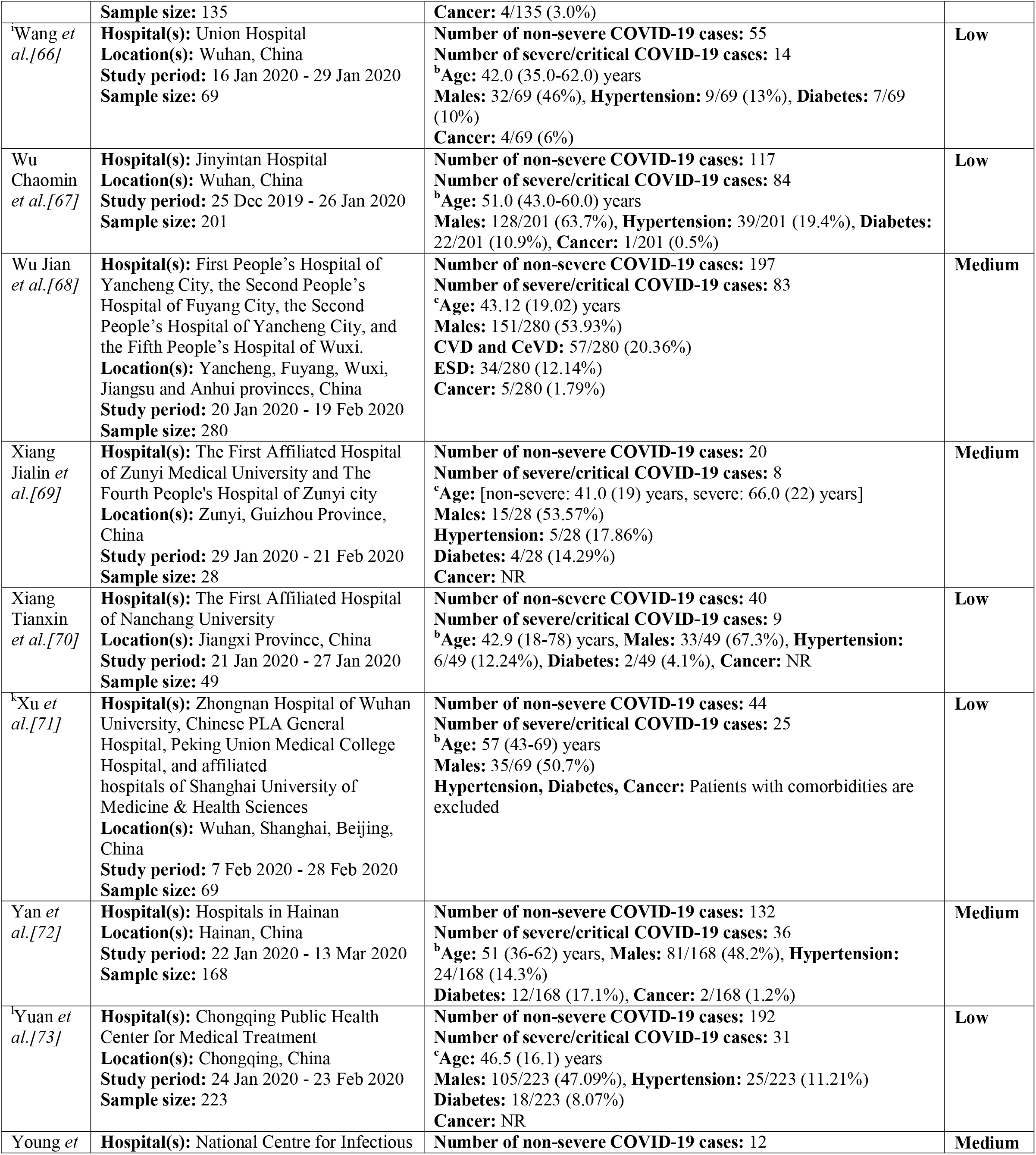
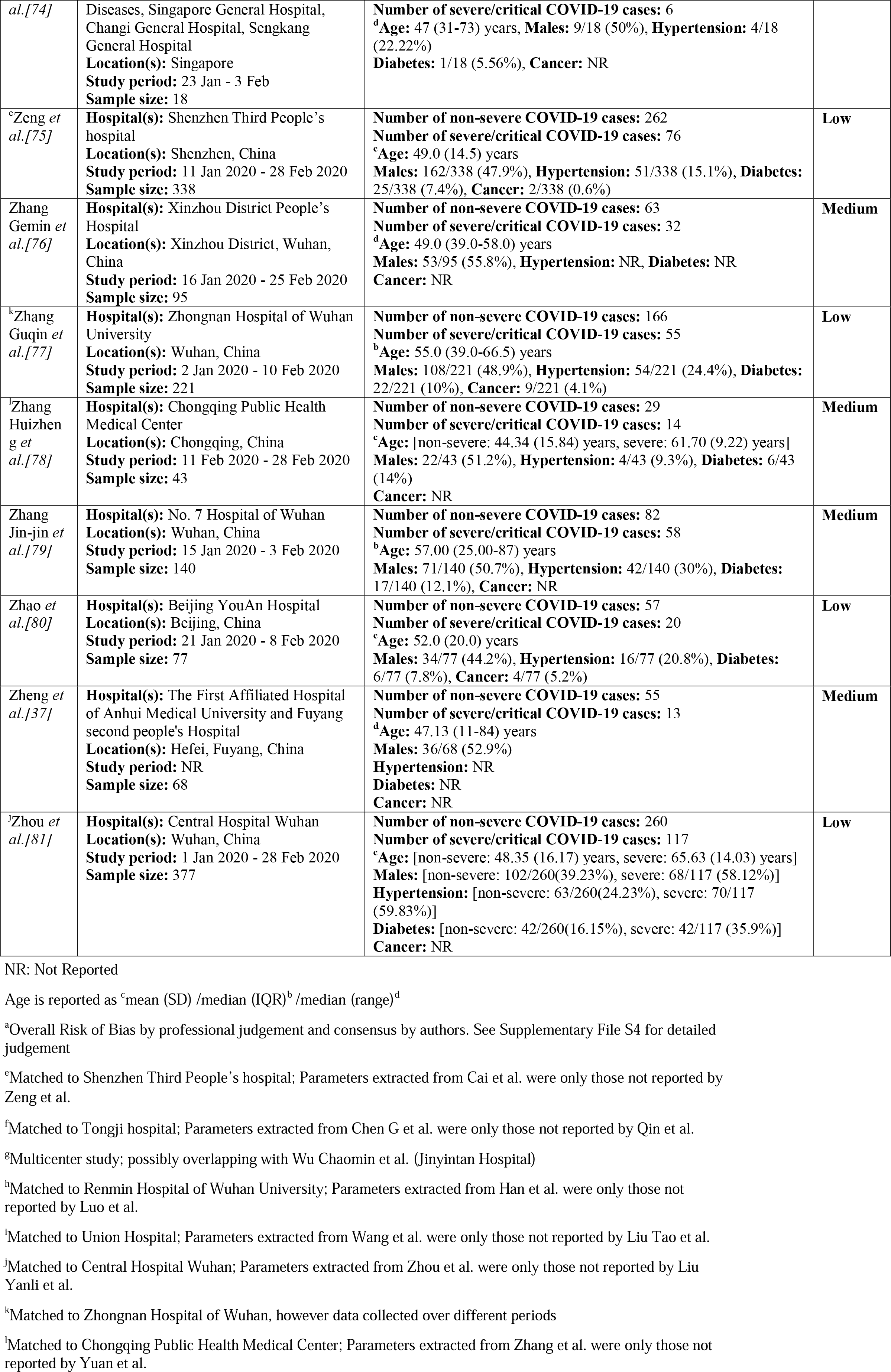
Characteristics of included studies

The median (or mean) age of patients in the included studies ranged from 35 years to 67 years, and the proportion of male patients ranged from 30% to 81%. Patients in the included studies had varying proportions of comorbidities such as hypertension, diabetes and cancer as detailed in Table 1 (and Supplement S3).

### Synthesis of results

Results of meta-analyses are reported in Table 2, and Forest plots and Leave-One-Out analysis are displayed in supplement S4.

**Table 2:** **Meta-estimates for severe or critical COVID-19 compared to non-severe COVID-19**

| Parameter | Number of studies | Number of persons | Meta estimate (95% CI) | p value | Prediction interval | I<sup>2</sup> | Q test p-value |
| --- | --- | --- | --- | --- | --- | --- | --- |
| **Hematological parameters** |  |  |  |  |  |  |  |
| **White cell count** |  |  |  |  |  |  |  |
| Continuous, MMD ($\times 10^9/L$) | 28 | 4749 | 0.87 (0.35, 1.40) | 0.001 | -1.54, 3.30 | 80.5% | <0.001 |
| *Without outlier studies* | 23 |  | 0.72 (0.36, 1.07) | <0.001 | -0.46, 1.89 | 45.8% | 0.009 |
| Leukocytosis, MPR | 11 | 3455 | 3.95 (2.35, 6.65) | <0.001 | 0.86, 18.22 | 64.3% | 0.002 |
| *Without outlier studies* | 10 |  | 3.21 (2.13, 4.82) | <0.001 | 1.18, 8.65 | 41.5% | 0.081 |
| **Neutrophils** |  |  |  |  |  |  |  |
| Continuous, MMD ($\times 10^9/L$) | 21 | 3091 | 1.23 (0.58, 1.88) | 0.001 | -1.59, 4.05 | 89.9% | <0.001 |
| *Without outlier studies* | 19 |  | 1.07 (0.71, 1.44) | <0.001 | -0.04, 2.18 | 46.0% | 0.015 |
| Neutrophilia, MPR | 6 | 1237 | 4.29 (1.74, 10.64) | 0.002 | 0.22, 84.80 | 85.6% | <0.001 |
| **Lymphocytes** |  |  |  |  |  |  |  |
| Continuous, MMD ($\times 10^9/L$) | 27 | 6465 | -0.38 (-0.46, -0.30) | <0.001 | -0.77, 0.01 | 84.0% | <0.001 |
| *Without outlier studies* | 24 |  | -0.38 (-0.45, -0.31) | <0.001 | -0.64, -0.12 | 57.3% | 0.001 |
| Lymphocytopenia, MPR | 14 | 3875 | 1.74 (1.43, 2.12) | <0.001 | 0.88, 3.42 | 92.5% | <0.001 |
| *Without outlier studies* | 9 |  | 1.85 (1.47, 2.33) | <0.001 | 0.97, 3.56 | 64.0% | 0.005 |
| **Monocytes** |  |  |  |  |  |  |  |
| Continuous, MMD ($\times 10^9/L$) | 14 | 2002 | -0.03 (-0.06, 0.01) | 0.102 | -0.14, 0.08 | 57.2% | 0.004 |
| *Without outlier studies* | 13 |  | -0.02 (-0.05, 0.01) | 0.188 | -0.11, 0.06 | 44.6% | 0.041 |
| **Platelets** |  |  |  |  |  |  |  |
| Continuous, MMD ($\times 10^9/L$) | 24 | 3877 | -21.47 (-41.12, -1.83) | 0.032 | -114.89, 71.94 | 92.0% | <0.001 |
| *Without outlier studies* | 23 |  | -18.95 (-28.52, -9.39) | <0.001 | -50.30, 12.38 | 47.9% | 0.006 |
| Thrombocytopenia, MPR | 10 | 2421 | 1.79 (1.30, 2.48) | <0.001 | 0.81, 3.98 | 48.4% | 0.042 |
| **Hemoglobin** |  |  |  |  |  |  |  |
| Continuous, MMD (g/dl) | 17 | 2931 | -0.33 (-0.57, -0.08) | 0.010 | -0.95, 0.30 | 32.5% | 0.096 |
| *Without outlier studies* | 16 |  | -0.31 (-0.53, -0.09) | 0.005 | -0.79, 0.17 | 21.2% | 0.212 |
| **CD3 count** |  |  |  |  |  |  |  |
| Continuous, MMD (cells/μl) | 6 | 601 | -380.82 (-515.30, -246.36) | <0.001 | -835.46, 73.80 | 80.1% | <0.001 |
| **CD4 count** |  |  |  |  |  |  |  |
| Continuous, MMD (cells/μl) | 7 | 669 | -204.86(-302.63, -107.10) | <0.001 | -539.07, 129.35 | 86.8% | <0.001 |
| **CD8 count** |  |  |  |  |  |  |  |
| Continuous, MMD (cells/μl) | 6 | 600 | -123.63 (-170.64, -76.61) | <0.001 | -270.44, 23.19 | 66.2% | 0.011 |
| **NLR** |  |  |  |  |  |  |  |
| Continuous, MMD | 5 | 1377 | 2.71 (1.82, 3.61) | <0.001 | -0.44, 5.87) | 79.7% | <0.001 |
| **SII** |  |  |  |  |  |  |  |
| Continuous, MMD | 2 | 487 | 394.00 (38.11, 749.87) | 0.030 | - | 84.6% | 0.010 |
| **Infection/inflammation-related indices** |  |  |  |  |  |  |  |
| **C-reactive protein (CRP)** |  |  |  |  |  |  |  |
| Continuous, MMD (mg/L) | 26 | 4959 | 38.62 (29.16, 48.07) | <0.001 | -6.01, 83.23 | 88.4% | <0.001 |
| *Without outlier studies* | 21 |  | 36.95 (29.30, 44.60) | <0.001 | 9.36, 64.53 | 59.8% | <0.001 |
| Elevated CRP, MPR | 13 | 2740 | 1.60 (1.32, 1.93) | <0.001 | 0.78, 3.27 | 93.4% | <0.001 |
| *Without outlier studies* | 11 |  | 1.59 (1.42, 1.77) | <0.001 | 1.18, 2.13 | 53.9% | 0.021 |
| **Erythrocyte sedimentation rate (ESR)** |  |  |  |  |  |  |  |
| Continuous, MMD (mm/hr) | 8 | 1705 | 20.01 (10.14, 29.87) | <0.001 | -13.27, 53.28 | 86.4% | <0.001 |
| *Without outlier studies* | 7 |  | *15.4 (7.14, 23.73)* | *<0.001* | *-11.09, 41.96* | *79.4%* | *<0.001* |
| Elevated ESR, MPR | 3 | 545 | 1.67 (0.67, 4.17) | 0.271 | 0.00, >100 | 97.9% | <0.001 |
| **Interleukin-6 (IL6)** |  |  |  |  |  |  |  |
| Continuous, MMD (pg/ml) | 7 | 1183 | 17.37 (4.74, 30.00) | 0.007 | -24.70, 59.56 | 94.7% | <0.001 |
| *Without outlier studies* | 6 |  | *20.61 (9.88, 31.33)* | *<0.001* | *-13.45, 54.67* | *81.4%* | *<0.001* |
| Elevated IL6, MPR | 3 | 357 | 2.15 (0.94, 5.00) | 0.067 | 0.00, >100 | 87.4% | <0.001 |
| *Without outlier studies* | 2 |  | *1.33 (1.07, 1.66)* | *0.001* | *-* | *0.0%* | *0.770* |
| **Procalcitonin (PCT)** |  |  |  |  |  |  |  |
| Continuous, MMD (ng/ml) | 18 | 4225 | 0.06 (0.04, 0.08) | <0.001 | -0.03, 0.15 | 89.5% | <0.001 |
| *Without outlier studies* | 13 |  | *0.05 (0.04, 0.06)* | *<0.001* | *0.03, 0.07* | *9.8%* | *0.348* |
| Elevated PCT, MPR | 12 | 2311 | 2.48 (1.78, 3.43) | <0.001 | 0.99, 6.19 | 53.6% | 0.014 |
| **Liver function parameters** |  |  |  |  |  |  |  |
| **Alanine aminotransferase (ALT)** |  |  |  |  |  |  |  |
| Continuous, MMD (U/L) | 25 | 4450 | 6.53 (4.43, 8.93) | <0.001 | 1.09, 11.97 | 25.4% | 0.122 |
| *Without outlier studies* | 21 |  | *5.21 (3.68, 6.73)* | *<0.001* | *3.59, 6.82* | *0.0%* | *0.754* |
| Elevated ALT, MPR | 12 | 2540 | 1.59 (1.36, 1.87) | <0.001 | 1.21, 2.09 | 10.3% | 0.344 |
| **Aspartate aminotransferase (AST)** |  |  |  |  |  |  |  |
| Continuous, MMD (U/L) | 25 | 4320 | 11.95 (8.80, 15.11) | <0.001 | -0.04, 23.95 | 68.8% | <0.001 |
| *Without outlier studies* | 22 |  | *10.63 (7.06, 14.19)* | *<0.001* | *7.06, 14.19* | *11.8%* | *0.302* |
| Elevated AST, MPR | 14 | 2705 | 2.14 (1.80, 2.54) | <0.001 | 1.43, 3.21 | 29.5% | 0.141 |
| **Total bilirubin** |  |  |  |  |  |  |  |
| Continuous, MMD (μmol/l) | 18 | 2104 | 1.62 (0.87, 2.37) | <0.001 | 0.81, 2.43 | 0.0% | 0.490 |
| Hyperbilirubinemia, MPR | 5 | 1704 | 1.70 (1.23, 2.35) | 0.001 | 1.01, 2.87 | 0.0% | 0.699 |
| **Total protein** |  |  |  |  |  |  |  |
| Continuous, MMD (g/L) | 5 | 482 | -1.49 (-3.19, 0.20) | 0.085 | -4.24, 1.26 | 0.0% | 0.763 |
| Hypoproteinemia, MPR | 2 | 208 | 1.65 (1.33, 2.04) | <0.001 | - | 0.0% | 0.658 |
| **Albumin** |  |  |  |  |  |  |  |
| Continuous, MMD (g/L) | 21 | 2891 | -4.58 (-6.21, -2.94) | <0.001 | -11.95, 2.79 | 95.4% | <0.001 |
| *Without outlier studies* | 19 |  | *-4.27 (-5.21, -3.33)* | *<0.001* | *-7.69, -0.85* | *67.9%* | *<0.001* |
| Hypoalbuminemia, MPR | 4 | 554 | 2.23 (1.93, 2.93) | <0.001 | 1.50, 3.77 | 0.0% | 0.726 |
| **Prealbumin** |  |  |  |  |  |  |  |
| Continuous, MMD (mg/dl) | 3 | 367 | -40.14 (-52.95, -27.33) | <0.001 | -133.59, 53.31 | 6.9% | 0.342 |
| **Globulin** |  |  |  |  |  |  |  |
| Continuous, MMD (g/L) | 4 | 476 | 2.31 (0.58, 4.04) | 0.009 | -4.61, 9.22 | 63.2% | 0.043 |
| *Without outlier studies* | 3 |  | *1.31 (0.30, 2.32)* | *0.011* | *-5.26, 7.88* | *0.0%* | *0.452* |
| **Kidney function parameters** |  |  |  |  |  |  |  |
| **Blood urea** |  |  |  |  |  |  |  |
| Continuous, MMD (mmol/l) | 19 | 2623 | 1.02 (0.66, 1.38) | <0.001 | -0.13, 2.17) | 46.1% | 0.015 |
| *Without outlier studies* | 18 |  | *1.09 (0.76, 1.42)* | *<0.001* | *0.16, 2.02* | *36.1%* | *0.064* |
| Elevated blood urea, MPR | 3 | 624 | 3.63 (1.73, 7.65) | <0.001 | 0.01, >100 | 39.9% | 0.189 |
| **Creatinine** |  |  |  |  |  |  |  |
| Continuous, MMD (μmol/l) | 26 | 4467 | 5.57 (3.12, 8.03) | <0.001 | -0.43, 11.57 | 18.7% | 0.197 |
| Elevated creatinine, MPR | 8 | 2019 | 1.90 (1.07, 3.36) | 0.027 | 0.48, 7.43 | 40.5% | 0.108 |
| **Cystatin C** |  |  |  |  |  |  |  |
| Continuous, MMD (mg/l) | 4 | 426 | 0.20 (0.10, 0.29) | <0.001 | -0.16, 0.55 | 52.8% | 0.095 |
| **Myocardial biomarkers** |  |  |  |  |  |  |  |
| **Creatine kinase muscle-brain** |  |  |  |  |  |  |  |
| Continuous, MMD (U/L) | 10 | 1324 | 1.48 (0.36, 2.59) | 0.009 | -0.75, 3.71 | 19.6% | 0.263 |
| Continuous, MMD (ng/ml) | 3 | 293 | 0.67 (0.19, 1.15) | 0.007 | -2.47, 3.18 | 0.0% | 0.964 |
| **Troponin I** |  |  |  |  |  |  |  |
| Continuous, MMD (ng/ml) | 8 | 2379 | 0.02 (0.00, 0.04) | 0.038 | -0.03, 0.08 | 79.7% | <0.001 |
| Elevated Troponin I | 3 | 831 | 4.00 (1.22, 13.2) | 0.022 | 0.00, >100 | 85.8% | 0.001 |
| **α-hydroxybutyric dehydrogenase** |  |  |  |  |  |  |  |
| Continuous, MMD (U/L) | 6 | 465 | 89.17 (45.26, 133.08) | <0.001 | -46.20, 224.55 | 67.6% | 0.009 |
| **Other biochemical parameters** |  |  |  |  |  |  |  |
| **Glucose** |  |  |  |  |  |  |  |
| Continuous, MMD (mmol/L) | 7 | 1343 | 1.02 (0.64, 1.39) | <0.001 | 0.20, 1.83 | 26.8% | 0.224 |
| Elevated glucose, MPR | 2 | 491 | 1.40 (1.15, 1.72) | 0.001 | - | 0.0% | 0.411 |
| **Cholinesterase** |  |  |  |  |  |  |  |
| Continuous, MMD (U/ml) | 2 | 229 | -1.11 (-1.79, -0.45) | 0.001 | - | 0.0% | 0.941 |
| **Lactate dehydrogenase (LDH)** |  |  |  |  |  |  |  |
| Continuous, MMD (U/L) | 22 | 2297 | 122.76 (94.14, 151.39) | <0.001 | 8.83, 236.70 | 72.6% | <0.001 |
| *Without outlier studies* | 20 |  | 114 (91.60, 138.21) | <0.001 | 39.84, 189.97 | 49.6% | 0.006 |
| Elevated LDH, MPR | 10 | 1893 | 2.41 (1.65, 3.51) | <0.001 | 0.70, 8.34 | 87.7% | <0.001 |
| *Without outlier studies* | 8 |  | 2.35 (1.65, 3.35) | <0.001 | 0.77, 7.17 | 85.7% | <0.001 |
| **Serum ferritin** |  |  |  |  |  |  |  |
| Continuous, MMD (μg/L) | 5 | 2342 | 430.28 (289.12, 571.45) | <0.001 | -5.40, 865.97 | 61.6% | <0.001 |
| Elevated ferritin, MPR | 2 | 412 | 2.3 (1.67, 3.17) | <0.001 | - | 0.0% | 0.511 |
| **Serum electrolytes** |  |  |  |  |  |  |  |
| **Sodium** |  |  |  |  |  |  |  |
| Continuous, MMD (mmol/L) | 10 | 1503 | -1.67 (-2.60, -0.74) | 0.001 | -3.98, 0.64 | 43.0% | 0.072 |
| **Potassium** |  |  |  |  |  |  |  |
| Continuous, MMD (mmol/L) | 12 | 1790 | -0.19 (-0.30, -0.10) | <0.001 | -0.46, 0.07 | 50.8% | 0.022 |
| Reduced potassium, MPR | 3 | 667 | 1.70 (1.14, 2.54) | 0.010 | 0.02, >100 | 58.5% | 0.090 |
| **Chloride** |  |  |  |  |  |  |  |
| Continuous, MMD (mmol/L) | 6 | 1074 | -1.49 (-3.08, 0.09) | 0.065 | -6.01, 3.03 | 57.5% | 0.038 |
| **Calcium** |  |  |  |  |  |  |  |
| Continuous, MMD (mmol/L) | 5 | 486 | -0.13 (-0.18, -0.09) | <0.001 | -0.26, -0.01 | 40.8% | 0.149 |
| **Coagulation parameters** |  |  |  |  |  |  |  |
| **Prothrombin time** |  |  |  |  |  |  |  |
| Continuous, MMD (s) | 16 | 1650 | 0.39 (0.14, 0.64) | 0.002 | -0.44, 1.22 | 68.2% | <0.001 |
| *Without outlier studies* | 15 |  | 0.29 (0.09, 0.48) | 0.004 | -0.22, 0.79 | 39.4% | 0.058 |
| **Activated partial thromboplastin** |  |  |  |  |  |  |  |
| Continuous, MMD (s) | 14 | 1918 | -0.49 (-1.95, 0.97) | 0.509 | -5.70, 4.72 | 77.6% | <0.001 |
| *Without outlier studies* | 12 |  | -0.33 (-1.50, 0.83) | 0.575 | -3.78, 3.11 | 55.2% | 0.011 |
| **D-dimer** |  |  |  |  |  |  |  |
| Continuous, MMD (mg/L) | 23 | 4740 | 0.52 (0.37, 0.66) | <0.001 | -0.02, 1.05 | 82.4% | <0.001 |
| *Without outlier studies* | 16 |  | 0.36 (0.27, 0.44) | <0.001 | 0.27, 0.45 | 0.0% | 0.561 |
| Elevated D-Dimer, MPR | 9 | 2030 | 2.27 (1.67, 3.09) | <0.001 | 0.87, 5.92 | 76.9% | <0.001 |
| *Without outlier studies* | 7 |  | 2.14 (1.81, 2.52) | <0.001 | 1.72, 2.65 | 0.0% | 0.435 |

*A study was considered an outlier if the study's confidence interval did not overlap with the confidence interval of the pooled effect*
MMD: meta-median difference; MPR: meta-prevalence ratio; NLR: neutrophil-to-lymphocyte ratio; SII: systemic inflammation Index; CRP: C-reactive protein; ESR: erythrocyte sedimentation rate; IL-6: interleukin-6; PCT: procalcitonin; ALT: alanine aminotransferase; AST: aspartate aminotransferase; LDH: lactate dehydrogenase

As pertains to haematological parameters, the majority of studies reported higher white cell count and higher neutrophil count in severe or critical COVID-19 patients relative to non-severe COVID-19 patients. Median difference in individual studies ranged from -1.6 to 7.3 (×10^9^ cells/L) for white cell count and from -1.0 to 5.2 (×10^9^ cells/L) for neutrophil count. The MMD estimates (10^9^ cells/L) were 0.87 (95% CI: 0.35 to 1.40; I^2^: 80.5%) for white cell count and 1.23 (95% CI 0.58 to 1.88; I^2^: 90%) for neutrophil count. When the results were expressed in terms of ratio measures, patients with severe or critical COVID-19 had significantly higher likelihood of having leucocytosis (MPR: 3.95 [95% CI: 2.35, 6.65], I^2^: 64%) and neutrophilia (MPR: 4.29 [95% CI: 1.74, 10.64], I^2^: 86%). All but one of 27 studies reported lower lymphocyte count in severe or critical COVID-19 patients relative to patients with non-severe disease. Median difference in individual studies ranged from -0.8 to 0.2 (×10^9^ cells/L). The MMD for lymphocyte count (×10^9^ cells/L) was -0.39 (95% -0.47, -0.31; I^2^: 78%), and the MPR for lymphopenia was 2.02 (95% CI: 1.52, 2.69; I^2^: 92%). Also, severe or critical COVID-19 patients had relatively lower CD3 count (MMD: -380.8 [-515.3, -246.4], I^2^: 80%), CD4 count (MMD: - 204.9 [-302.6, -107.1], I^2^: 87%) and CD8 count (MMD: -123.6 [-170.6, – 76.6] I^2^: 66%); all differences measured in terms of cells/μl.

All studies that examined data on inflammation indices reported higher CRP, ESR and IL-6 level in severely or critically ill patients. Median difference in individual studies ranged from 8.1 to 83.3 mg/L for CRP, from 4.7 to 52.4 mm/hr for ESR, and from 1.1 to 101.4 pg/ml for IL-6. The corresponding MMD (95% CI; I^2^) estimates were 36.97 (27.58, 46.35; 85%), 21.93 (10.59, 33.28; 88% for ESR, and 17.37 (4.74, 30.00; 95%) for IL-6. The MPR values for elevated CRP, ESR and IL-6 were 1.50 [95% CI: 1.26, 1.77; I2: 91%), 1.67 (95% CI: 0.67, 4.18; I2: 98%) and 2.15 (95% CI: 0.95, 4.90; I^2^: 87%), respectively, although the data for the last two parameters were limited to just three studies. Higher levels of ferritin, a positive acute-phase reactant, were positively associated with severe or critical COVID-19 (MMD: 451.86 μg/L [95% CI: 212.91, 690.82] I^2^: 71%), whereas the same association with albumin, a negative acute-phase reactant, was in the opposite direction (MMD: -4.99 g/L [95% CI: -6.47, -3.51], I^2^: 87%).

Additional significant differences between patients with severe or critical COVID-19 and their non-severely ill counterparts were observed for liver enzymes, ALT (MMD: 6.89 U/L [95% CI; 4.69, 9.10], I^2^: 17%) and AST (MMD: 11.96 U/L [95% CI: 8.56, 15.37] I^2^: 68%); kidney function parameters, urea (MMD: 1.04 mmol/l [95% CI: 0.64, 1.45], I^2^: 48%) and creatinine (MMD: 4.87 μmol/l [95% CI: 2.40, 7.35], I^2^: 7%); biomarkers of myocardial function, troponin I (MMD: 0.01 ng/ml [95% CI: 0.00, 0.02], I^2^: 0%) and CK-MB (MMD: 1.46 U/L [95% CI:0.22, 2.70], I^2^: 28%); measures of coagulation, D-dimer (MMD: 0.65 mg/ml [95% CI: 0.45, 0.85], I^2^: 84%) and platelet count (MMD: -21.48 ×10^9^ cells/L [95% CI: -41.12, -1.83], I^2^: 92%); and lactate dehydrogenase, a marker of tissue damage (MMD: 124.26 U/L [95% CI: 92.89, 155.64], I^2^: 74%).

### Assessment of threats to validity

The threats to validity in this meta-analysis fall into two broad categories: risk of bias in individual studies, and publication bias across the body of literature. Assessments of these two categories of threat to validity are presented below.

Using the NIH study quality assessment tools, 28 studies (62.2%) were rated as having a low risk of bias, 14 studies (31.1%) were rated as having a medium risk of bias, and 3 studies (6.7%) were rated as having a high risk of bias. The majority of studies had a clearly defined study objective (97.8%), a well-defined study population (100%), and had comparable subjects (100%). In contrast, no study provided a sample size calculation or power description. All the studies were rated as having a high risk of bias for the element assessing a temporal sequence between the laboratory measure and disease severity, and none of the reported results was adjusted for potential confounding (Figure 2 and supplement S5). Our results did not markedly differ in sensitivity analyses after excluding studies with a high risk of bias.

**Figure 2:**
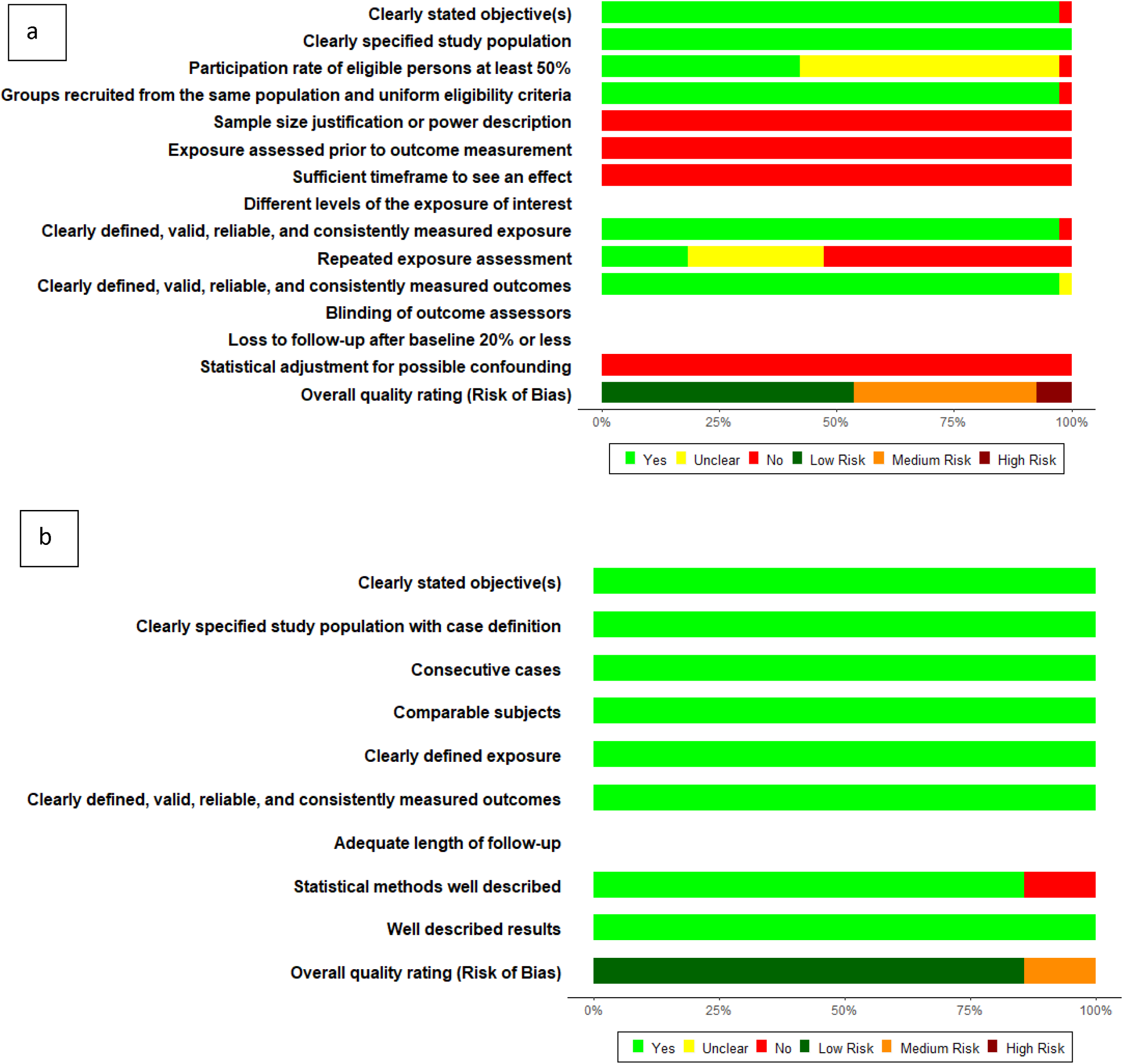
Summary plot for Risk of Bias assessment; A: Risk of bias assessment for 38 retrospective cohort/cross sectional studies; B: Risk of bias assessment for 7 case series studies

The symmetry of funnel plots obtained from the 4 laboratory parameters with the highest number of individual studies was assessed using Egger’s test. The symmetrical funnel plots for C-reactive protein (*p*: 0.155) and creatinine (*p*: 0.415) suggested no evidence of publication bias whereas asymmetrical funnel plot for white cell count (*p*: 0.004) and lymphocyte count (*p*: 0.005) indicated significant influence of smaller studies, which may be indicative of publication bias (Figure 3). Important to note that Egger’s test may not be robust for C-reactive protein, white cell count and lymphocyte parameters due to substantial heterogeneity (I^2^>75%).

**Figure 3:**
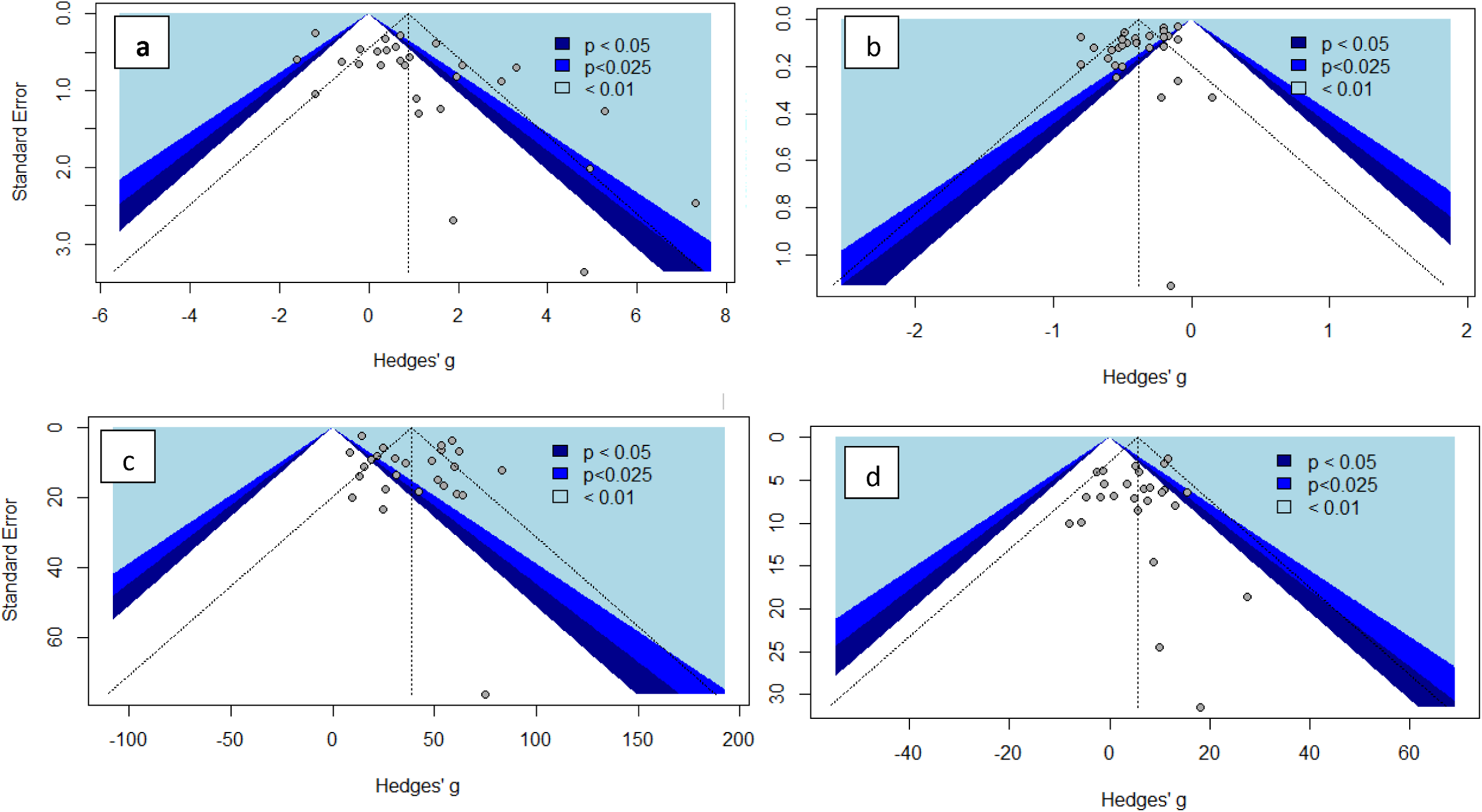
Funnel Plots: a: White cell count, Egger’s test, p=0.004; b: Lymphocytes, Egger’s test, p= 0.005; c : C-Reactive protein, Egger’s test, p=0.155; d: Creatinine, Egger’s test, p= 0.415

### Additional analysis

In sensitivity analyses excluding outlier studies, statistical heterogeneity was reduced, and the meta-estimate of most laboratory parameters were not markedly altered. In sensitivity analysis using mean differences (supplement S6), there was substantial heterogeneity for most laboratory parameters, and the associations observed from using median differences persisted.

## Discussion

COVID-19 is a rapidly evolving pandemic with significant global morbidity and mortality. The aim of this meta-analysis was to investigate which clinical laboratory parameters may be associated with severe or critical COVID-19 disease. Out of the 39 clinical laboratory parameters evaluated, we found that derangements in 36 clinical laboratory parameters were significantly associated with severe or critical COVID-19. Whilst some of the observed associations may not be clinically relevant, certain, more pronounced laboratory abnormalities may have important clinical implications. Markers of an overactive innate immune system such as markedly elevated neutrophil-to-lymphocyte ratio (NLR), IL-6, serum ferritin and C-reactive protein, and markers of a deficient adaptive immune system such as lymphocytes and CD4 count could help recognise potential severe infections during triage, while markers of organ failure could be helpful in monitoring evolution of hospitalised COVID-19 patients.

Following infection with a virus, the innate immune system in activated. This early response is nonspecific and serves to limit virus multiplication during the acute phase [33]. The adaptive immune system is activated a few days later and is responsible for a more specific response, which is immunomodulatory (via engagement of helper T cells and regulatory T cells) and produces ‘immunological memory’ [33]. Elevated lymphocyte count is commonly found in most viral infections, and the magnitude and quality of T cell responses may determine the fate of these infections [34, 35]. Failure to mount an appropriate adaptive immune response means the innate immune response remains continuously stimulated with deleterious effects on the lungs and other organs. We found that severe or critical COVID-19 patients had increased markers of innate immune system activity compared to patients with non-severe disease. This is evidenced in the significantly higher levels of neutrophils, IL-6, and acute phase reaction markers such as CRP, ESR and serum ferritin, as well as decreased concentrations of albumin and prealbumin. Severe or critical COVID-19 patients also exhibited defective adaptive immune response evidenced by significantly lower levels of lymphocytes and their subsets (CD3, CD4, CD8). CD4 count is currently being used to define severe cases of HIV infection [36]. In the case of HIV, the virus directly infects CD4 cells using the envelope glycoprotein gp120. Various authors have suggested that SARS-CoV-2 could deplete lymphocytes directly by infecting T lymphocytes, or indirectly through lymphocyte apoptosis induced by persistent elevated inflammatory cytokines [15, 37, 38]. Since severe COVID-19 patients display reduced lymphocyte count, it is likely that the cytokine release syndrome observed in some patients with severe or critical COVID-19 is mediated by interferons, TNFs, and cytokines secreted by non-T cell leucocytes such as macrophages, neutrophils and NK cells which are all key elements of innate immunity to viruses [39].

These findings could be applied clinically to identify severe or critical COVID-19 patients. For example, routine monitoring of NLR may provide insight into the functioning of both the innate and adaptive immune responses and help predict the clinical course of COVID-19. Despite the 45 studies included in this review, only five reported results for NLR; all these five studies found a significant association between increased NLR and severe or critical COVID-19 disease.

We also found that patients with severe or critical COVID-19 had significantly higher biomarkers of tissue and organ damage such as LDH, liver enzymes, kidney function parameters and markers of myocardial function. These observed associations could be explained by 3 mechanisms. First, the virus may cause direct organ damage by attaching to the ACE2 receptors, which are commonly expressed in the lungs, heart, arteries, kidneys and intestines [40]. The second, more indirect mechanism is systemic hyperinflammation caused by the cytokine release syndrome mediated by the innate immune system [40]. Systemic hyperinflammation affects all organs and could also explain the significantly increased expression of markers of disseminated intravascular coagulation (high D-dimer and depleted platelet count) in severely or critically ill patients [41, 42]. The third, also indirect, mechanism by which severe or critical COVID-19 causes multiple organ damage is hypoxia resulting from respiratory failure.

Once the mechanisms of COVID-19 induced organ damage are better understood, markers reflecting the pathophysiological changes caused by these mechanisms may find their way into clinical practice. Based on the results of our meta-analysis especially promising may be markers of immune function such as NLR, IL-6, C-reactive protein, serum ferritin, lymphocytes, CD4 count, and markers of coagulation and organ damage such as D-dimer, LDH, troponin I and liver enzymes.

The strengths and limitations of this review and meta-analysis need to be considered in the context of rapidly evolving literature. On the one hand, our study identified some associations that deserve further consideration and may lead to improvements in the risk stratification, monitoring and management of COVID-19 patients. On the other hand, it is important to emphasize that our analyses need to be viewed as hypothesis-generating rather than hypothesis-testing. Due to the large number of associations examined simultaneously there is a considerable likelihood of false-positive findings. This limitation can be addressed in future, more focused, studies that will take into consideration prior knowledge and reduce the likelihood of false-positive results through application of Bayesian and empirical-Bayes methods [43]. Our review is also affected by the limitations of the underlying literature. Of those, perhaps the most important is the cross-sectional nature of the analyses used in most publications. Although it is plausible that markers of immune function can be used to predict disease severity, the evidence would have been stronger if the studies were able to perform laboratory testing of COVID-19 patients before their disease severity was known. In addition, many studies from China reported on overlapping patient populations. While we tried to exclude studies that relied on the same data, it is possible that some of the associations examined in this meta-analysis were based on non-independent observations.

## Conclusions

Compared to non-severe COVID-19, severe or critical COVID-19 is associated with increased markers of innate immune response such as neutrophil count, NLR, IL-6, CRP and serum ferritin; decreased markers of adaptive immune response such as lymphocyte, CD4 and CD8 counts; and increased markers of tissue damage and major organ failure including D-dimer LDH, Troponin I, CK-MB, AST, ALT, urea, and creatinine. Based on the results of our meta-analysis, especially promising markers are NLR, IL-6, serum ferritin, lymphocyte and CD4 counts, D-dimer and troponin I. The clinical value of these markers should be explored further to assess the risk of severe or critical disease and to monitor the clinical course of COVID-19.

## Data Availability

All data used in this review are available from the individual studies

## Acronyms and Abbreviations

ACE2: Angiotensin-converting enzyme 2
ALT: Alanine aminotransferase
AST: Aspartate aminotransferase
CI: Confidence interval
CK-MB: Creatine kinase muscle-brain
COVID-19: Coronavirus disease 2019
CRP: C-reactive protein
ESR: Erythrocyte sedimentation rate
IL-6: Interleukin-6
IQR: Interquartile range
LDH: Lactate dehydrogenase
MERS-CoV: Middle East Respiratory Syndrome Coronavirus
MMD: Meta-median difference
MOOSE: Meta-analysis of Observational Studies in Epidemiology
MPR: Meta-prevalence ratios
NIH: National Institutes of Health
NLR: Neutrophil-to-lymphocyte ratio
PRISMA: Preferred Reporting Items for Systematic Reviews and Meta-Analyses
PROSPERO: International Prospective Register of Systematic Reviews
SARS-CoV: Severe Acute Respiratory Syndrome Coronavirus
SARS-CoV-2: Severe Acute Respiratory Syndrome Coronavirus 2
SD: Standard deviation

## Article Information

### Author Contributions

JM and PP had full access to all the data in the study and take responsibility for the integrity of the data and the accuracy of the data analysis.

### Study concept and design

JM, PP, AK, MN

### Acquisition, analysis, or interpretation of data

JM, PP, SU, KM

### Drafting of the manuscript

SU, KM

### Critical revision of the manuscript for important intellectual content

MG, AK, MN

### Statistical analysis

JM

### Study supervision

MG

## References

1. Gorbalenya AE, Baker SC, Baric RS, de Groot RJ, Drosten C, Gulyaeva AA, et al. The species Severe acute respiratory syndrome-related coronavirus: classifying 2019-nCoV and naming it SARS-CoV-2. Nature Microbiology. 2020;5(4):536–44. doi: 10.1038/s41564-020-0695-z.

2. Zhu N, Zhang D, Wang W, Li X, Yang B, Song J, et al. A Novel Coronavirus from Patients with Pneumonia in China, 2019. N Engl J Med. 2020;382(8):727-33. Epub 2020/01/25. doi: 10.1056/NEJMoa2001017. PubMed PMID: 31978945; PubMed Central PMCID: PMCPMC7092803.

3. Lu R, Zhao X, Li J, Niu P, Yang B, Wu H, et al. Genomic characterisation and epidemiology of 2019 novel coronavirus: implications for virus origins and receptor binding. Lancet. 2020;395(10224):565-74. Epub 2020/02/03. doi: 10.1016/s0140-6736(20)30251-8. PubMed PMID: 32007145.

4. Huang C, Wang Y, Li X, Ren L, Zhao J, Hu Y, et al. Clinical features of patients infected with 2019 novel coronavirus in Wuhan, China. Lancet. 2020;395(10223):497-506. PubMed PMID: 31986264.

5. Li Q, Guan X, Wu P, Wang X, Zhou L, Tong Y, et al. Early Transmission Dynamics in Wuhan, China, of Novel Coronavirus-Infected Pneumonia. The New England journal of medicine. 2020;382(13):1199-207. Epub 2020/01/29. doi: 10.1056/NEJMoa2001316. PubMed PMID: 31995857.

6. Cucinotta D, Vanelli M. WHO Declares COVID-19 a Pandemic. Acta Biomed. 2020;91(1):157-60. Epub 2020/03/20. doi: 10.23750/abm.v91i1.9397. PubMed PMID: 32191675.

7. Dong E, Du H, Gardner L. An interactive web-based dashboard to track COVID-19 in real time. The Lancet Infectious Diseases. doi: 10.1016/S1473-3099(20)30120-1.

8. Wu Z, McGoogan JM. Characteristics of and Important Lessons From the Coronavirus Disease 2019 (COVID-19) Outbreak in China: Summary of a Report of 72lJ314 Cases From the Chinese Center for Disease Control and Prevention. JAMA. 2020;323(13):1239–42. doi: 10.1001/jama.2020.2648.

9. Cascella M, Rajnik M, Cuomo A, Dulebohn SC, Di Napoli R. Features, Evaluation and Treatment Coronavirus (COVID-19). StatPearls Publishing. 2020;01:01. PubMed PMID: 32150360.

10. Rodriguez-Morales AJ, Cardona-Ospina JA, Gutiérrez-Ocampo E, Villamizar-Peña R, Holguin-Rivera Y, Escalera-Antezana JP, et al. Clinical, laboratory and imaging features of COVID-19: A systematic review and meta-analysis. Travel Medicine and Infectious Disease. 2020:101623. doi: https://doi.org/10.1016/j.tmaid.2020.101623.

11. Weiss P, Murdoch DR. Clinical course and mortality risk of severe COVID-19. Lancet. 2020;395(10229):1014-5. PubMed PMID: 32197108.

12. Yang X, Yu Y, Xu J, Shu H, Xia J, Liu H, et al. Clinical course and outcomes of critically ill patients with SARS-CoV-2 pneumonia in Wuhan, China: a single-centered, retrospective, observational study. The Lancet Respiratory Medicine. 2020;24:24. PubMed PMID: 32105632.

13. Guan WJ, Ni ZY, Hu Y, Liang WH, Ou CQ, He JX, et al. Clinical Characteristics of Coronavirus Disease 2019 in China. New England Journal of Medicine. 2020;28:28. PubMed PMID: 32109013.

14. Herold T, Jurinovic V, Arnreich C, Hellmuth JC, Bergwelt-Baildon M, Klein M, et al. Level of IL-6 predicts respiratory failure in hospitalized symptomatic COVID-19 patients. medRxiv. 2020:2020.04.01.20047381. doi: 10.1101/2020.04.01.20047381.

15. Tan L, Wang Q, Zhang D, Ding J, Huang Q, Tang Y-Q, et al. Lymphopenia predicts disease severity of COVID-19: a descriptive and predictive study. medRxiv. 2020:2020.03.01.20029074. doi: 10.1101/2020.03.01.20029074.

16. Tabata S, Imai K, Kawano S, Ikeda M, Kodama T, Miyoshi K, et al. Non-severe vs severe symptomatic COVID-19: 104 cases from the outbreak on the cruise ship “Diamond Princess” in Japan. medRxiv. 2020:2020.03.18.20038125. doi: 10.1101/2020.03.18.20038125.

17. Moutchia J, Pokharel P, Kerri A, McGaw K, Uchai S, Nji MAM. Clinical laboratory parameters associated with severe or critical novel coronavirus disease 2019 (COVID-19): a systematic review and meta-analysis of observational studies: PROSPERO; 2020 [cited 2020 Apr 17]. Available from: https://www.crd.york.ac.uk/prospero/display_record.php?RecordID=176651.

18. Moher D, Liberati A, Tetzlaff J, Altman DG, The PG. Preferred Reporting Items for Systematic Reviews and Meta-Analyses: The PRISMA Statement. PLOS Medicine. 2009;6(7):e1000097. doi: 10.1371/journal.pmed.1000097.

19. Stroup DF, Berlin JA, Morton SC, Olkin I, Williamson GD, Rennie D, et al. Meta-analysis of observational studies in epidemiology: a proposal for reporting. Meta-analysis Of Observational Studies in Epidemiology (MOOSE) group. Jama. 2000;283(15):2008-12. Epub 2000/05/02. doi: 10.1001/jama.283.15.2008. PubMed PMID: 10789670.

20. World Health Organisation. Clinical management of severe acute respiratory infection (SARI) when COVID-19 disease is suspected (interim guidance) 2020 [cited 2020 17 Apr]. Available from: https://apps.who.int/iris/bitstream/handle/10665/331446/WHO-2019-nCoV-clinical-2020.4-eng.pdf?sequence=1&IsAllowed=y.

21. National Health Commission of the People’s Republic of China. Diagnosis and Treatment Protocol for Novel Coronavirus Pneumonia (Trial version 7) 2020 [cited 2020 17 Apr]. Available from: http://busan.china-consulate.org/chn/zt/4/P020200310548447287942.pdf.

22. China National Health Commission. Diagnosis and Treatment Protocol for Novel Coronavirus Pneumonia (Trial version 7) 2020 [cited 2020 17 Apr]. Available from: http://busan.china-consulate.org/chn/zt/4/P020200310548447287942.pdf.

23. Covidence. Better systematic review management 2020 [cited 2020 Apr 1]. Available from: https://www.covidence.org/home.

24. National Institutes of Health. Study Quality Assessment Tools 2020 [cited 2020 Apr 1]. Available from: https://www.nhlbi.nih.gov/health-topics/study-quality-assessment-tools.

25. Harrer M, Cuijpers P, Furukawa TA, Ebert DD. Doing Meta-Analysis in R: A Hands-on Guide 2019 [cited 2020 Apr 16]. Available from: https://bookdown.org/MathiasHarrer/Doing_Meta_Analysis_in_R/.

26. McGrath S, Sohn H, Steele R, Benedetti A. Meta-analysis of the difference of medians. Biometrical Journal. 2020;62(1):69–98. doi: 10.1002/bimj.201900036.

27. Wan X, Wang W, Liu J, Tong T. Estimating the sample mean and standard deviation from the sample size, median, range and/or interquartile range. BMC Med Res Methodol. 2014;14:135. Epub 2014/12/20. doi: 10.1186/1471-2288-14-135. PubMed PMID: 25524443; PubMed Central PMCID: PMCPMC4383202.

28. Luo D, Wan X, Liu J, Tong T. Optimally estimating the sample mean from the sample size, median, mid-range, and/or mid-quartile range. Stat Methods Med Res. 2018;27(6):1785-805. Epub 2016/09/30. doi: 10.1177/0962280216669183. PubMed PMID: 27683581.

29. The R Foundation. The R Project for Statistical Computing [cited 2020 Apr 17]. Available from: https://www.r-project.org/.

30. Liu J, Liu Y, Xiang P, Pu L, Xiong H, Li C, et al. Neutrophil-to-Lymphocyte Ratio Predicts Severe Illness Patients with 2019 Novel Coronavirus in the Early Stage. medRxiv. 2020:2020.02.10.20021584. doi: 10.1101/2020.02.10.20021584.

31. Wu C, Chen X, Cai Y, Xia J, Xu S, Huang H, et al. Risk Factors Associated with Acute Respiratory Distress Syndrome and Death in Patients with Coronavirus Disease 2019 Pneumonia in Wuhan, China. JAMA Internal Medicine. 2020. PubMed PMID: 631235378.

32. Dai Z, Gao L, Luo D, Xiao J, Huang C, Zeng G, et al. [Analysis of clinical characteristics of new coronavirus pneumonia in Hunan Province]. Practical Preventive Medicine. 2020:1–4.

33. Janeway C. Immunobiology : the immune system in health and disease. 6th ed ed. New York: Garland Science; 2005.

34. Kamphorst AO, Ahmed R. CD4 T-cell immunotherapy for chronic viral infections and cancer. Immunotherapy. 2013;5(9):975–87. doi: 10.2217/imt.13.91. PubMed PMID: 23998732.

35. Naeim F, Rao PN, Grody WW. Chapter 19 - Non-neoplastic and Borderline Lymphocytic Disorders. In:Naeim F, Rao PN, Grody WW, editors. Hematopathology. Oxford: Academic Press; 2008. p. 455–76.

36. Kagan JM, Sanchez AM, Landay A, Denny TN. A Brief Chronicle of CD4 as a Biomarker for HIV/AIDS: A Tribute to the Memory of John L. Fahey. For Immunopathol Dis Therap. 2015;6(1-2):55- doi: 10.1615/ForumImmunDisTher.2016014169. PubMed PMID: 27182452.

37. Zheng M, Gao Y, Wang G, Song G, Liu S, Sun D, et al. Functional exhaustion of antiviral lymphocytes in COVID-19 patients. Cellular & Molecular Immunology. 2020;19:19. PubMed PMID: 32203188.

38. Wang X, Xu W, Hu G, Xia S, Sun Z, Liu Z, et al. SARS-CoV-2 infects T lymphocytes through its spike protein-mediated membrane fusion. Cellular & Molecular Immunology. 2020;07:07. PubMed PMID: 32265513.

39. Tisoncik JR, Korth MJ, Simmons CP, Farrar J, Martin TR, Katze MG. Into the eye of the cytokine storm. Microbiol Mol Biol Rev. 2012;76(1):16–32. doi: 10.1128/MMBR.05015-11. PubMed PMID: 22390970.

40. Shi Y, Wang Y, Shao C, Huang J, Gan J, Huang X, et al. COVID-19 infection: the perspectives on immune responses. Cell Death & Differentiation. 2020. doi: 10.1038/s41418-020-0530-3.

41. Han H, Yang L, Liu R, Liu F, Wu KL, Li J, et al. Prominent changes in blood coagulation of patients with SARS-CoV-2 infection. Clinical Chemistry & Laboratory Medicine. 2020;16:16. PubMed PMID: 32172226.

42. Wang J, Hajizadeh N, Moore EE, McIntyre RC, Moore PK, Veress LA, et al. Tissue Plasminogen Activator (tPA) Treatment for COVID-19 Associated Acute Respiratory Distress Syndrome (ARDS): A Case Series. J Thromb Haemost. 2020. Epub 2020/04/09. doi: 10.1111/jth.14828. PubMed PMID: 32267998.

43. Hamra GB, MacLehose RF, Cole SR. Sensitivity analyses for sparse-data problems-using weakly informative bayesian priors. Epidemiology. 2013;24(2):233-9. Epub 2013/01/23. doi: 10.1097/EDE.0b013e318280db1d. PubMed PMID: 23337241; PubMed Central PMCID: PMCPMC3607322.

44. Cai Q, Huang D, Ou P, Yu H, Zhu Z, Xia Z, et al. COVID-19 in a designated infectious diseases hospital outside Hubei Province, China. Allergy. 2020;n/a(n/a). doi: 10.1111/all.14309.

45. Cao M, Zhang D, Wang Y, Lu Y, Zhu X, Li Y, et al. Clinical Features of Patients Infected with the 2019 Novel Coronavirus (COVID-19) in Shanghai, China. medRxiv. 2020:2020.03.04.20030395. doi: 10.1101/2020.03.04.20030395.

46. Cao W. Clinical features and laboratory inspection of novel coronavirus pneumonia (COVID-19) in Xiangyang, Hubei. medRxiv. 2020:2020.02.23.20026963. doi: 10.1101/2020.02.23.20026963.

47. chen d, Li X, song q, Hu C, Su F, Dai J. Hypokalemia and Clinical Implications in Patients with Coronavirus Disease 2019 (COVID-19). medRxiv. 2020:2020.02.27.20028530. doi: 10.1101/2020.02.27.20028530.

48. Chen G, Wu D, Guo W, Cao Y, Huang D, Wang H, et al. Clinical and immunologic features in severe and moderate Coronavirus Disease 2019. The Journal of clinical investigation. 2020;27. PubMed PMID: 631358781.

49. chen m, tu c, Tan C, Zheng X, wang x, wu j, et al. Key to successful treatment of COVID-19: accurate identification of severe risks and early intervention of disease progression. medRxiv. 2020:2020.04.06.20054890. doi: 10.1101/2020.04.06.20054890.

50. Fang X, Mei Q, Yang T, Zhang L, Yang Y, Wang Y, et al. [2019 New Coronavirus Infected Pneumonia: Clinical Features and Treatment Analysis of 79 Cases]. Chinese Pharmacology Bulletin. 2020;36(4):1–7.

51. Gong J, Ou J, Qiu X, Jie Y, Chen Y, Yuan L, et al. A Tool to Early Predict Severe 2019-Novel Coronavirus Pneumonia (COVID-19) : A Multicenter Study using the Risk Nomogram in Wuhan and Guangdong, China. medRxiv. 2020:2020.03.17.20037515. doi: 10.1101/2020.03.17.20037515.

52. Goyal P, Choi JJ, Pinheiro LC, Schenck EJ, Chen R, Jabri A, et al. Clinical Characteristics of Covid-19 in New York City. The New England journal of medicine. 2020. doi: https://dx.doi.org/10.1056/NEJMc2010419.

53. Han Y, Zhang H, Mu S, Wei W, Jin C, Xue Y, et al. Lactate dehydrogenase, a Risk Factor of Severe COVID-19 Patients. medRxiv. 2020:2020.03.24.20040162. doi: 10.1101/2020.03.24.20040162.

54. Hu L, Chen S, Fu Y, Gao Z, Long H, Ren H-w, et al. Risk Factors Associated with Clinical Outcomes in 323 COVID-19 Patients in Wuhan, China. medRxiv. 2020:2020.03.25.20037721. doi: 10.1101/2020.03.25.20037721.

55. Lescure FX, Bouadma L, Nguyen D, Parisey M, Wicky PH, Behillil S, et al. Clinical and virological data of the first cases of COVID-19 in Europe: a case series. The Lancet Infectious Diseases. 2020;27:27. PubMed PMID: 32224310.

56. Liu C, Jiang ZC, Shao CX, Zhang HG, Yue HM, Chen ZH, et al. Preliminary study of the relationship between novel coronavirus pneumonia and liver function damage: a multicenter study. [Chinese]. Zhonghua gan zang bing za zhi = Zhonghua ganzangbing zazhi = Chinese journal of hepatology. 2020;28(2):148-52. PubMed PMID: 631012069.

57. Liu M, He P, Liu HG, Wang XJ, Li FJ, Chen S, et al. [Clinical characteristics of 30 medical workers infected with new coronavirus pneumonia]. Chung-Hua Chieh Ho Ho Hu Hsi Tsa Chih Chinese Journal of Tuberculosis & Respiratory Diseases. 2020;43(3):209-14. PubMed PMID: 32164090.

58. Liu T, Zhang J, Yang Y, Ma H, Li Z, Zhang J, et al. The potential role of IL-6 in monitoring severe case of coronavirus disease 2019. medRxiv. 2020:2020.03.01.20029769. doi: 10.1101/2020.03.01.20029769.

59. Liu Y, Sun W, Li J, Chen L, Wang Y, Zhang L, et al. Clinical features and progression of acute respiratory distress syndrome in coronavirus disease 2019. medRxiv. 2020:2020.02.17.20024166. doi: 10.1101/2020.02.17.20024166.

60. Luo X, Zhou W, Yan X, Guo T, Wang B, Xia H, et al. Prognostic value of C-reactive protein in patients with COVID-19. medRxiv. 2020:2020.03.21.20040360. doi: 10.1101/2020.03.21.20040360.

61. Petrilli CM, Jones SA, Yang J, Rajagopalan H, Donnell LF, Chernyak Y, et al. Factors associated with hospitalization and critical illness among 4,103 patients with COVID-19 disease in New York City. medRxiv. 2020:2020.04.08.20057794. doi: 10.1101/2020.04.08.20057794.

62. Qian GQ, Yang NB, Ding F, Ma AHY, Shen YF, Shi CW, et al. Epidemiologic and Clinical Characteristics of 91 Hospitalized Patients with COVID-19 in Zhejiang, China: A retrospective, multicentre case series. QJM : monthly journal of the Association of Physicians. 2020;17. PubMed PMID: 631268529.

63. Qin C, Zhou L, Hu Z, Zhang S, Yang S, Tao Y, et al. Dysregulation of immune response in patients with COVID-19 in Wuhan, China. Clinical Infectious Diseases. 2020;12:12. PubMed PMID: 32161940.

64. Qu R, Ling Y, Zhang YH, Wei LY, Chen X, Li XM, et al. Platelet-to-lymphocyte ratio is associated with prognosis in patients with coronavirus disease-19. Journal of Medical Virology. 2020;17:17. PubMed PMID: 32181903.

65. Wan S, Xiang Y, Fang W, Zheng Y, Li B, Hu Y, et al. Clinical Features and Treatment of COVID-19 Patients in Northeast Chongqing. Journal of Medical Virology. 2020;21:21. PubMed PMID: 32198776.

66. Wang Z, Yang B, Li Q, Wen L, Zhang R. Clinical Features of 69 Cases with Coronavirus Disease 2019 in Wuhan, China. Clinical Infectious Diseases. 2020;16:16. PubMed PMID: 32176772.

67. Wu C, Chen X, Cai Y, Xia J, Zhou X, Xu S, et al. Risk Factors Associated With Acute Respiratory Distress Syndrome and Death in Patients With Coronavirus Disease 2019 Pneumonia in Wuhan, China. JAMA Internal Medicine. 2020;13:13. PubMed PMID: 32167524.

68. Wu J, Li W, Shi X, Chen Z, Jiang B, Liu J, et al. Early antiviral treatment contributes to alleviate the severity and improve the prognosis of patients with novel coronavirus disease (COVID-19). Journal of Internal Medicine. 2020;27:27. PubMed PMID: 32220033.

69. Xiang J, Wen J, Yuan X, Xiong S, Zhou XUE, Liu C, et al. Potential biochemical markers to identify severe cases among COVID-19 patients. medRxiv. 2020:2020.03.19.20034447. doi: 10.1101/2020.03.19.20034447.

70. Xiang T, Liu J, Xu F, Cheng N, Liu Y, Qian K, et al. [Clinical characteristics of 49 patients with novel coronavirus pneumonia in Jiangxi area]. Chinese Journal of Respiratory and Critical Care Medicine. 2020:1–7.

71. Xu Y, Li Y-r, Zeng Q, Lu Z-b, Li Y-z, Wu W, et al. Clinical Characteristics of SARS-CoV-2 Pneumonia Compared to Controls in Chinese Han Population. medRxiv. 2020:2020.03.08.20031658. doi: 10.1101/2020.03.08.20031658.

72. Yan S, Song X, Lin F, Zhu H, Wang X, Li M, et al. Clinical Characteristics of Coronavirus Disease 2019 in Hainan, China. medRxiv. 2020:2020.03.19.20038539. doi: 10.1101/2020.03.19.20038539.

73. Yuan J, Sun Y, Zuo Y, Chen T, Cao Q, Yuan G, et al. [Clinical characteristics of 223 patients with new coronavirus pneumonia in Chongqing]. Journal of Southwest University (Natural Science Edition). 2020:1–7.

74. Young BE, Ong SWX, Kalimuddin S, Low JG, Tan SY, Loh J, et al. Epidemiologic Features and Clinical Course of Patients Infected With SARS-CoV-2 in Singapore. Jama. 2020;03:03. PubMed PMID: 32125362.

75. Zeng L, Li J, Liao M, Hua R, Huang P, Zhang M, et al. Risk assessment of progression to severe conditions for patients with COVID-19 pneumonia: a single-center retrospective study. medRxiv. 2020:2020.03.25.20043166. doi: 10.1101/2020.03.25.20043166.

76. Zhang G, Zhang J, Wang B, Zhu X, Wang Q, Qiu S. Analysis of clinical characteristics and laboratory findings of 95 cases of 2019 novel coronavirus pneumonia in Wuhan, China: a retrospective analysis. Respiratory Research. 2020;21(1):74. PubMed PMID: 32216803.

77. Zhang G, Hu C, Luo L, Fang F, Chen Y, Li J, et al. Clinical features and outcomes of 221 patients with COVID-19 in Wuhan, China. medRxiv. 2020:2020.03.02.20030452. doi: 10.1101/2020.03.02.20030452.

78. zhang h, wang x, fu z, luo m, zhang z, zhang k, et al. Potential Factors for Prediction of Disease Severity of COVID-19 Patients. medRxiv. 2020:2020.03.20.20039818. doi: 10.1101/2020.03.20.20039818.

79. Zhang JJ, Dong X, Cao YY, Yuan YD, Yang YB, Yan YQ, et al. Clinical characteristics of 140 patients infected with SARS-CoV-2 in Wuhan, China. Allergy. 2020;19:19. PubMed PMID: 32077115.

80. Zhao W, Yu S, Zha X, Wang N, Pang Q, Li T, et al. Clinical characteristics and durations of hospitalized patients with COVID-19 in Beijing: a retrospective cohort study. medRxiv. 2020:2020.03.13.20035436. doi: 10.1101/2020.03.13.20035436.

81. Zhou Y, Yang Z, Guo Y, Geng S, Gao S, Ye S, et al. A New Predictor of Disease Severity in Patients with COVID-19 in Wuhan, China. medRxiv. 2020:2020.03.24.20042119. doi: 10.1101/2020.03.24.20042119.

